# Aspartate/alanine aminotransferase ratio and development of chronic kidney disease in non-diabetic men and women: a population-based longitudinal study in Kagawa, Japan

**DOI:** 10.1101/2024.05.20.24307032

**Authors:** Yukari Okawa, Toshiharu Mitsuhashi, Etsuji Suzuki

## Abstract

**Background:** The relationship between serum aspartate/alanine aminotransferase ratio (AST/ALT) and subsequent development of chronic kidney disease (CKD) in non-diabetic Asian adults has not yet been fully investigated in longitudinal studies.

**Methods:** The study included all middle-aged and older non-diabetic Japanese citizens who received health check-ups in Zentsuji, Kagawa, Japan (1998–2023). AST/ALT was classified into three categories: <1.0 (reference), 1.0–<1.5, and ≥1.5. CKD was defined as an estimated glomerular filtration rate of <60 mL/min/1.73 m^2^. The Weibull accelerated failure time model was used to examine the association between AST/ALT categories and subsequent CKD onset because the proportional hazards assumption was violated.

**Results:** Of 6309 men and 9192 women, 2966 men and 4395 women remained in the final cohort. After a mean follow-up of 7.50 years for men and 8.34 years for women, 33.7% of men and 34.0% of women developed CKD. Women had higher AST/ALT than men. In women, a dose-response relationship was observed, with a 9% shorter survival time to CKD onset for AST/ALT ≥1.5 compared with AST/ALT <1.0. In contrast, men had a shorter survival time to CKD onset by point estimates, but the 95% confidence intervals crossed 1 in all models.

**Conclusions:** In this study comparing the risks of CKD development in non-diabetic men and women by AST/ALT levels, a dose-response relationship was only observed in women. Differences in the distribution of AST/ALT by sex may have affected the results. Therefore, in non-diabetic Japanese women, AST/ALT may be used as an indicator of future CKD development.

## Introduction

The serum aspartate/alanine aminotransferase ratio (AST/ALT), also called the De Ritis ratio, is a beneficial indicator for the aetiology of liver diseases[1]. AST/ALT is typically higher in women than in men, ≥2.0 if alcoholic liver disease is present, and <1.0 if chronic hepatitis or chronic viral hepatitis is present[1,2]. AST/ALT is also recognized as a risk factor for a variety of outcomes, with higher levels increasing the risk of the incidence of cancer, cardiovascular disease, and death[3–5]. However, the relationship between AST/ALT and chronic kidney disease (CKD) remains unclear.

CKD is a well-known public health problem, affecting 9.1% of the world’s population and causing 1.23 million deaths in 2017[6]. In general, CKD progresses gradually without symptoms until it reaches an irreversible state. If CKD remains untreated and progresses to end-stage renal disease, renal replacement therapies such as dialysis or kidney transplantation are required[7]. However, access to these expensive therapies is limited, especially in low-income countries, conferring physical, psychological, and financial burdens on patients[8]. Consequently, identification of risk factors for the early stages of CKD onset has important public health implications for preventing CKD and improving people’s quality of life.

A previous cross-sectional study investigated the relationship between AST/ALT and CKD prevalence in 29133 middle-aged Japanese women[9]. The study reported an adjusted odds ratio of 1.43 (95% confidence interval [CI]: 1.30–1.58) for the prevalence of CKD in those with AST/ALT <1 compared with those with AST/ALT ≥1, regardless of gamma-glutamyl transferase (GGT) elevation. However, because the study had a cross-sectional design and only included women, the findings only provided a snapshot of one female population at one point in time, and it was impossible to identify cause and effect. Moreover, the study included both non-diabetic and diabetic participants, although diabetes is a common cause of liver diseases and CKD[9–12]. Therefore, follow-up studies involving men and women without diabetes are needed to clarify the relationship between AST/ALT and new-onset CKD.

Accordingly, we aimed to evaluate the relationship between AST/ALT and CKD development in diabetes-free Japanese men and women in a longitudinal study.

## Materials and methods

### Data source

This study is an ongoing longitudinal study in Zentsuji, Kagawa Prefecture, Japan. Zentsuji City is located in the northeastern part of Shikoku Island, one of the four main islands of Japan. The city had a population of 30505 (49.6% men) on 1 April 2023. The city database includes data for annual health check-ups since 1998. We used the same database in previous studies[13,14]. In the present study, data were extracted from the database on 6 July 2023.

### Annual health check-ups

All citizens with fiscal year (FY) age ≥40 years are eligible to receive one health check-up per FY under their own volition. In FY1998 and FY1999, the city expanded the targeted citizens to include 35–39 years of age on a trial basis to promote the health of younger people. Approximately 30%–40% of the eligible population receive an annual health check-up on average.

The city follows the protocol of the Japanese Ministry of Health, Labour and Welfare when conducting the annual health check-ups[15]. Each check-up includes height and weight measurements, blood pressure testing, blood and urine tests, and a self-reported questionnaire about lifestyle habits such as alcohol intake and smoking.

### Study participants

This study aimed to evaluate the relationship between AST/ALT levels and subsequent development of CKD among non-diabetic Japanese men and women. Therefore, participants with non-Japanese nationality, haemoglobin A1C (HbA1c) ≥6.5% at study entry, glycosuria ≥1+ at study entry, or a single health check-up were excluded from the analysis. Participants with missing data for outcome or exposure variables, HbA1c values, or glycosuria levels at study entry were also excluded.

### Variables

#### Outcome

The outcome variable was development of CKD. The estimated glomerular filtration rate (eGFR) was used as a measure of renal function, with eGFR <60 mL/min/1.73 m^2^ defined as prevalent CKD[7]. The three-variable Japanese equation was used to calculate eGFR: 194 × *serum creatinine*(*mg/dL*)^−1.094^ × *age*(*years*)^−0.287^ (×0.739 *if female*)[16]. Serum creatinine (SCr) was measured to two decimal places using enzymatic methods.

#### Exposure

The exposure variable was AST/ALT levels. Serum AST (U/L) and ALT (U/L) were measured to zero decimal places using an ultraviolet spectrophotometric method. AST/ALT was classified into four categories: <1.0 (reference), 1.0–<1.5, 1.5–<2.0, and ≥2.0[9]. Owing to the small number of participants with AST/ALT ≥2.0, the AST/ALT ≥2.0 category was combined with the 1.5–<2.0 category.

#### Other covariates

In an attempt to reduce potential confounding bias, we adjusted for the following covariates: age category (34–59 [reference]/60–69/70–100), body mass index (BMI) category (normal weight [reference]/overweight or obesity), self-reported alcohol intake (non- or seldom-drinker [reference]/drinker), self-reported smoking status (non- or ex-smoker [reference]/smoker), hypertension (normal [reference]/hypertensive), dyslipidaemia (normal [reference]/dyslipidaemic), HbA1c value (%), and residential district (East [reference]/West/Central/South/Fudeoka/Tatsukawa/Yogita/Yoshiwara). The residential districts were adjusted to minimize the health impacts of neighbourhood characteristics[17].

BMI was calculated as weight in kilograms divided by height in meters squared, rounded to one decimal place, and dichotomized into normal weight (<25.0 kg/m^2^) and overweight or obesity (≥25.0 kg/m^2^)[18]. Hypertension was defined as systolic blood pressure ≥130 mmHg and/or diastolic blood pressure ≥80 mmHg[19]. Dyslipidaemia was regarded as serum low-density lipoprotein ≥140 mg/dL, serum high-density lipoprotein <40 mg/dL, and/or serum triglyceride ≥150 mg/dL[20]. On 1 April 2013, Japan changed the reporting units for HbA1c from the Japan Diabetes Society (JDS) unit (%) to the National Glycohaemoglobin Standardization Program (NGSP) unit (%). HbA1c levels in JDS units were converted to NGSP units using the following officially certified formula: 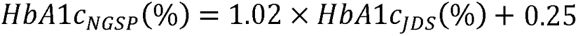[21].

### Statistical analysis

All analyses were stratified by sex. The participants’ demographic characteristics were summarized by the AST/ALT categories. The number of failures, person-years at risk, and incidence rate per 1000 person-years (IR) were displayed for binary or categorical covariates. The mean and standard deviation (SD) were shown for continuous covariates. Person-years were calculated from the date of study entry to the date of onset of CKD or diabetes, otherwise to the date of last check-up. Because this study aimed to examine the relationship between AST/ALT and CKD onset in a non-diabetic population, those who developed diabetes during the follow-up period were treated as right-censored. HbA1c ≥6.5% was defined as prevalent diabetes[22].

Kaplan–Meier curves by AST/ALT categories were generated for men and women. Violation of the proportional hazards (PH) assumption was confirmed by the Schoenfeld residuals and log–log plots. Thus, the Weibull accelerated failure time (AFT) model was selected according to the Akaike and Bayesian information criteria. The outcome measure of the Weibull AFT model was the time ratio; a higher time ratio indicates a longer survival time to CKD onset (e.g., a time ratio of 0.6 indicates a 40% shorter survival time to CKD onset compared with the reference category). Only estimates of exposure (i.e. AST/ALT) effects were presented to avoid the Table 2 fallacy.

In addition to the Crude model, four adjusted models were added to reduce confounding bias by reference to previous studies[9,23]: Model 1 was adjusted for age category; Model 2 was further adjusted for BMI category, self-reported alcohol intake, and self-reported smoking status; Model 3 was further adjusted for prevalent hypertension, dyslipidaemia, and HbA1c value; and Model 4 was further adjusted for residential district. In all analyses, a multiplicative term was added to the models if an interaction effect was observed between the exposure variable and any of the following covariates: sex, age category, BMI category, self-reported alcohol intake, self-reported smoking status, hypertension, dyslipidaemia, and HbA1c value.

A two-sided p-value <0.05 was considered statistically significant. All statistical analyses were conducted using Stata/MP 16.1 (StataCorp, College Station, TX, USA). A flow chart for the participants was created in Python 3.11.7. The district map of Zentsuji City was drawn in R 4.3.2. Geographic information system data were downloaded from the portal site of official statistics of Japan[24].

### Missing data

Participation in the health check-ups is voluntary. Consequently, there were missing values in this study. In the final cohort, the respective proportions of missing values for men and women were as follows: BMI category (0.02% and 0.02%), hypertension (0.01% and 0.03%), dyslipidaemia (16.7% and 24.2%), self-reported alcohol intake (31.2% and 28.6%), self-reported smoking status (25.1% and 23.1%), and residential district (1.32% and 1.43%).

Missing values were imputed using multiple imputation methods because we assumed that the pattern of missingness was missing at random. Binary variables were imputed by logistic regression and categorical variables by multinomial logistic regression[25]. The number of imputations was set to 40 times.

In this analysis, information on medication use (antihypertensive agents, lipid-lowering agents, and hypoglycaemic agents) could not be used because 58.9% of all observations were missing. There were some changes in the questionnaire issued by the Ministry of Health, Labour and Welfare, in that a question about medication use was added in 2012 but removed in 2020[15].

### Sensitivity analyses

Four sensitivity analyses were performed in this study. In the first sensitivity analysis, we used AST, AST, and GGT as exposure variables to determine which enzymes had the greatest impact on CKD development. ALT and AST were categorized as <30 (reference), 30–<40, and ≥40 U/L[9,26]. GGT was measured to zero decimal places using visible absorption spectroscopy and categorized as <30, 30–<50, and ≥50 U/L[9,27].

In general, eGFR values vary between observations. Therefore, the second sensitivity analysis used a more stringent definition of CKD to minimize the impact of eGFR variability leading to misclassification. Specifically, two consecutive observations of eGFR <60 mL/min/1.73 m^2^ were treated as CKD.

For the third sensitivity analysis, we defined CKD as dipstick proteinuria ≥1+, instead of eGFR <60 mL/min/1.73 m^2^, to confirm the effect of proteinuria on incident CKD. Finally, as the fourth sensitivity analysis, we used the Cox PH model, although the PH assumption was violated.

## Results

### Main analysis

In total, 6309 men and 9192 women agreed to participate in the study, with a mean age at study entry of 63.8 years for men and 61.3 years for women. After application of the exclusion criteria, 2966 men and 4395 women remained in the final cohort (Figure 1), with a mean age at study entry of 62.0 years (SD 10.6) for men and 59.2 years (SD 12.0) for women. The proportions of men and women who had CKD at study entry and were excluded from the analysis were 8.40% and 17.5% in the 34–59 age group, 25.8% and 31.8% in the 60–69 age group, and 47.7% and 54.3% in the 70–100 age group, respectively. At study entry, the mean AST/ALT values were 1.19 (SD 0.45) for men and 1.33 (SD 0.40) for women. The district map of Zentsuji is shown in Figure 2. The Kaplan–Meier survival estimates for men and women stratified by AST/ALT categories are shown in Figure 3.

**Figure 1.**
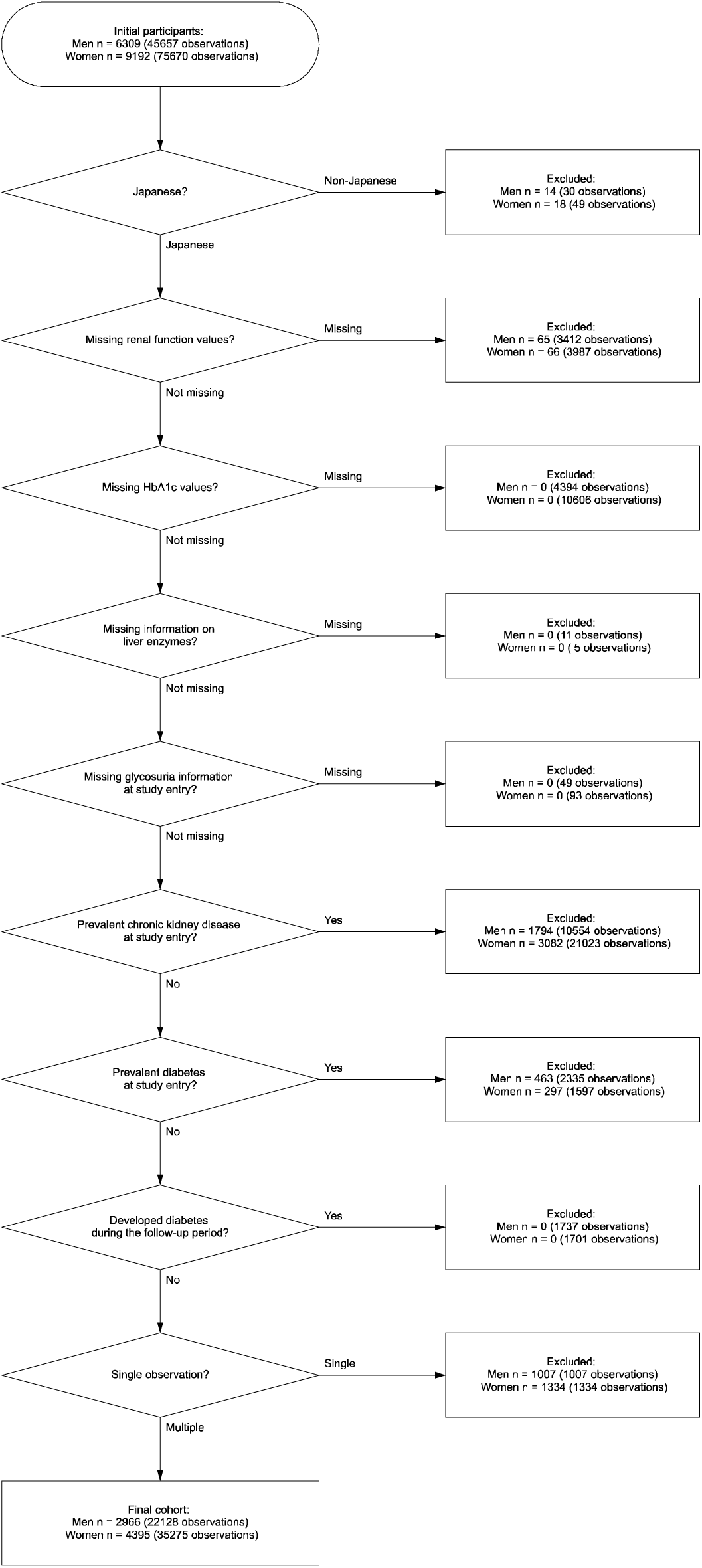
Flow chart of participants in the study cohort.

**Figure 2.**
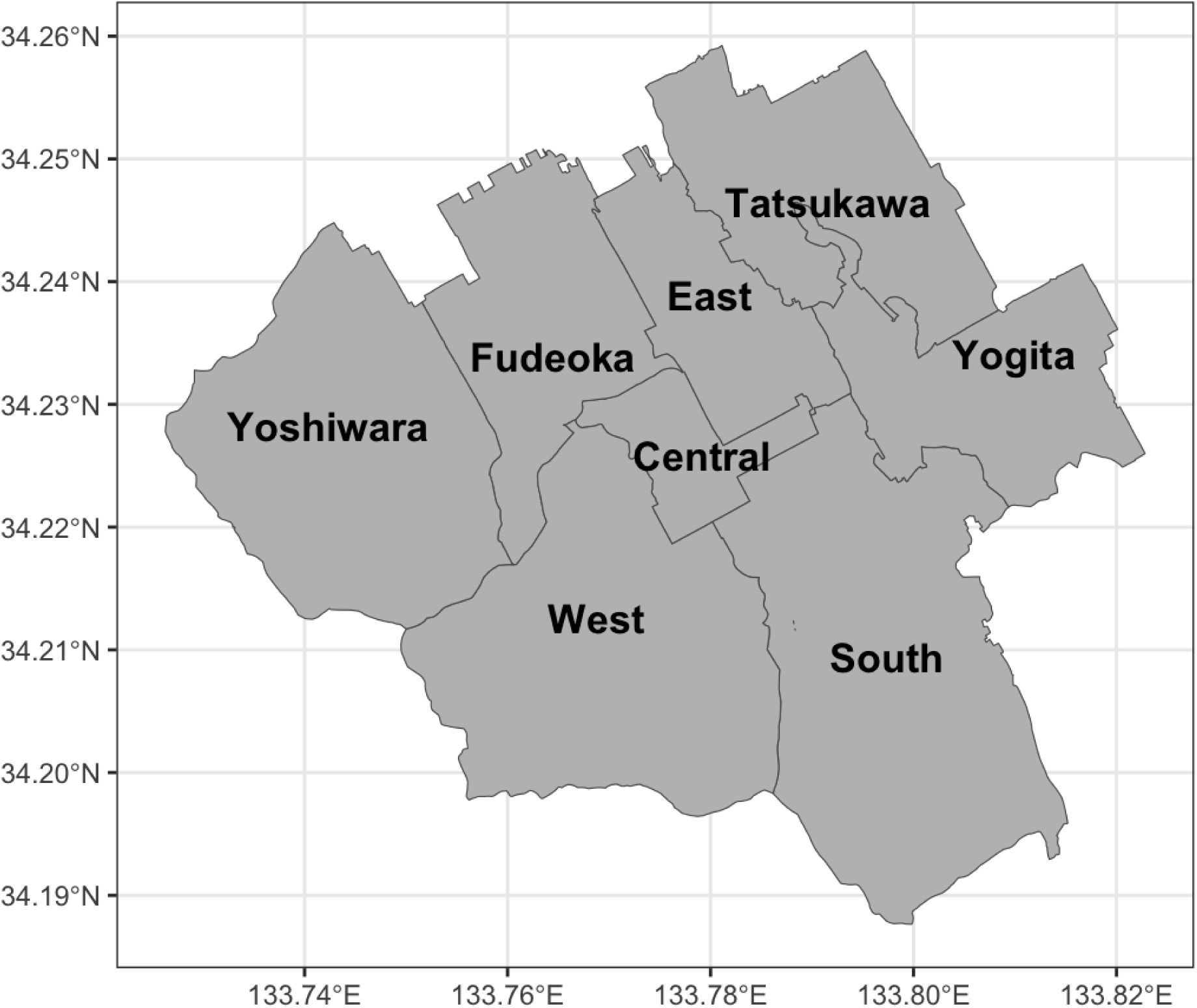
Map showing the districts in Zentsuji City.

**Figure 3.**
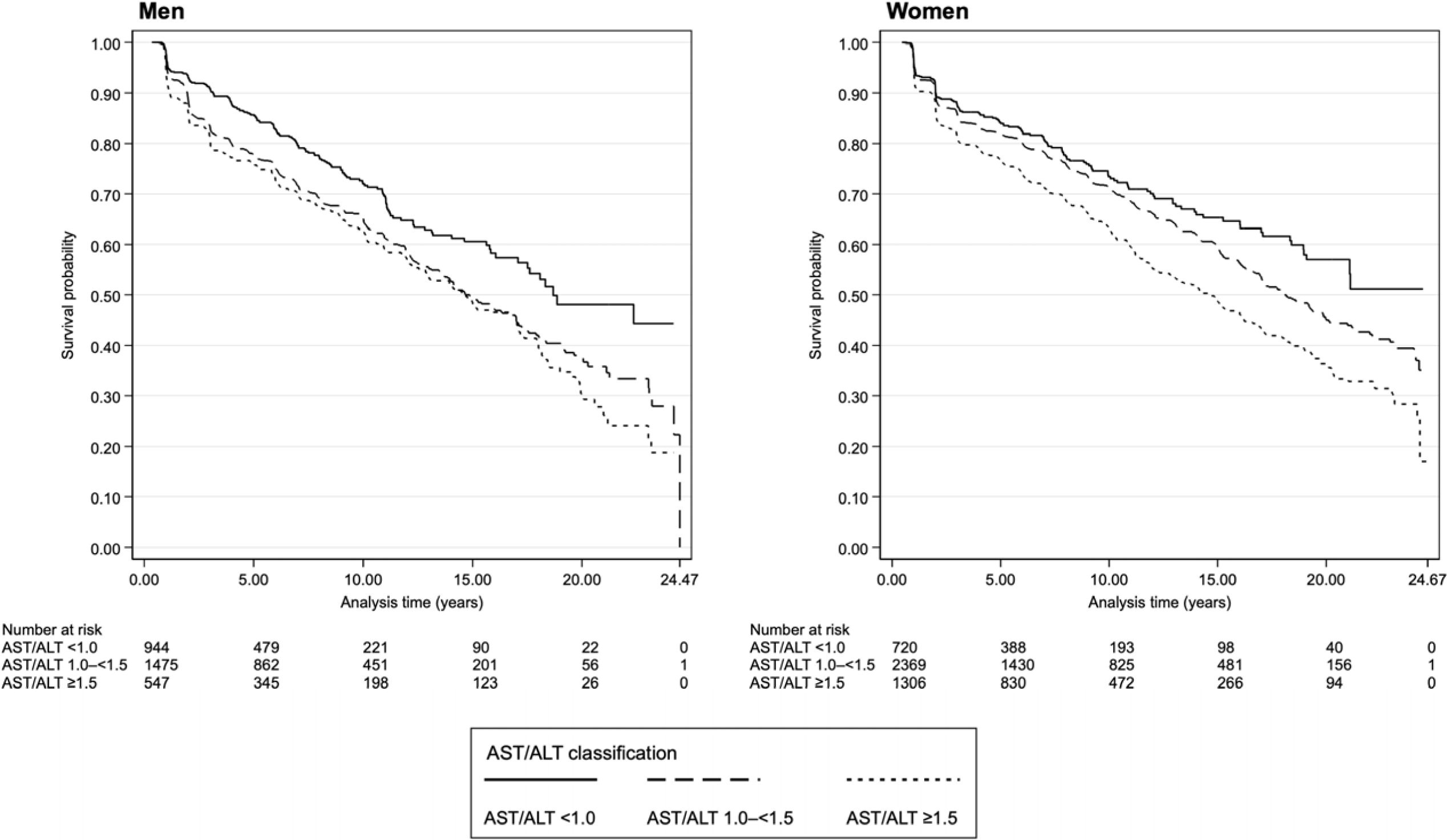
Kaplan–Meier survival estimates by AST/ALT categories for men and women.

Tables 1 and 2 show the participant characteristics for men and women, respectively. The mean follow-up period for women was slightly longer than that for men. After approximately 25 years of follow-up, 33.7% of men and 34.0% of women developed new-onset CKD. At CKD onset, men had a slightly older mean age (69.3 [SD 9.16] years) than women (67.5 [SD 11.2] years), and a lower mean AST/ALT value (1.30 [SD 0.50]) than women (1.42 [SD 0.43]). For both men and women, >95% of the CKD stage at CKD onset was G3a, the least severe stage. Specifically, the numbers (proportions of total) for stages G3a, G3b, G4, and G5 were 968 (96.8%), 27 (2.70%), 5 (0.50%), and 0 (0.00%) for men and 1429 (95.7%), 58 (3.88%), 3 (0.20%), and 4 (0.27%) for women, respectively. In both sexes, higher AST/ALT was associated with higher IR. Approximately half of the follow-up time belonged to the AST/ALT 1.0–<1.5 category regardless of sex. Men with higher AST/ALT tended to be normal weight, drinkers, and/or non-dyslipidaemic (Table 1). Women with higher AST/ALT tended to be normal weight, non-smokers, and/or non-dyslipidaemic (Table 2).

**Table 1.** Descriptive statistics of all observations stratified by AST/ALT ratio categories among 2966 non-diabetic Japanese male citizens of Zentsuji City (1998–2023)

|  | AST/ALT ratio classification |  |  |  |  |  |  |  |  | Total |  |  |
| --- | --- | --- | --- | --- | --- | --- | --- | --- | --- | --- | --- | --- |
|  | <1.0 |  |  | 1.0–<1.5 |  |  | ≥1.5 |  |  |  |  |  |
|  | Failure | PY | IR† | Failure | PY | IR† | Failure | PY | IR† | Failure | PY | IR† |
| Time at risk |  |  |  |  |  |  |  |  |  |  |  |  |
| Total |  | 6268.3 |  |  | 11164.3 |  |  | 4801.9 |  |  | 22234.5 |  |
| Minimum |  | 0.34 |  |  | 0.40 |  |  | 0.46 |  |  | 0.34 |  |
| Maximum |  | 24.2 |  |  | 24.5 |  |  | 22.4 |  |  | 24.5 |  |
| Mean |  | 4.64 |  |  | 5.22 |  |  | 4.54 |  |  | 7.50 |  |
| Median |  | 3.09 |  |  | 3.95 |  |  | 3.04 |  |  | 6.04 |  |
| Failure/IR† | 209 |  | 33.3 | 537 |  | 48.1 | 254 |  | 52.9 | 1000 |  | 45.0 |
| Variables |  |  |  |  |  |  |  |  |  |  |  |  |
| Age category in years |  |  |  |  |  |  |  |  |  |  |  |  |
| 34–59 | 42 | 2901.2 | 14.5 | 48 | 2681.6 | 17.9 | 12 | 588.3 | 20.4 | 102 | 6171.2 | 16.5 |
| 60–69 | 105 | 2289.9 | 45.9 | 218 | 4333.4 | 50.3 | 70 | 1526.5 | 45.9 | 393 | 8149.7 | 48.2 |
| 70–100 | 62 | 1077.2 | 57.6 | 271 | 4149.3 | 65.3 | 172 | 2687.2 | 64.0 | 505 | 7913.6 | 63.8 |
| BMI category‡ |  |  |  |  |  |  |  |  |  |  |  |  |
| Normal | 106 | 3550.8 | 29.9 | 394 | 8609.3 | 45.8 | 213 | 4249.6 | 50.1 | 713 | 16409.7 | 43.4 |
| Overweight or obesity | 103 | 2717.4 | 37.9 | 143 | 2555.0 | 56.0 | 41 | 552.3 | 74.2 | 287 | 5824.7 | 49.3 |
| Self-reported alcohol intake |  |  |  |  |  |  |  |  |  |  |  |  |
| Non- or seldom-drinker | 93.9 | 2580.8 | 36.4 | 210.9 | 4133.4 | 51.0 | 95.1 | 1687.6 | 56.4 | 399.9 | 8401.7 | 47.6 |
| Drinker | 115.1 | 3687.5 | 31.2 | 326.1 | 7030.9 | 46.4 | 158.9 | 3114.3 | 51.0 | 600.1 | 13832.7 | 43.4 |
| Self-reported smoking status |  |  |  |  |  |  |  |  |  |  |  |  |
| Non- or ex-smoker | 148.7 | 4182.3 | 35.6 | 402.3 | 7877.5 | 51.1 | 189.4 | 3339.7 | 56.7 | 740.4 | 15399.4 | 48.1 |
| Smoker | 60.3 | 2086.0 | 28.9 | 134.7 | 3286.8 | 41.0 | 64.6 | 1462.3 | 44.2 | 259.7 | 6835.1 | 38.0 |
| Hypertension§ |  |  |  |  |  |  |  |  |  |  |  |  |
| Normal | 81.0 | 2607.7 | 31.1 | 212.0 | 5105.5 | 41.5 | 92.0 | 2093.8 | 43.9 | 385.0 | 9807.1 | 39.3 |
| Hypertensive | 128.0 | 3660.5 | 35.0 | 325.0 | 6058.8 | 53.6 | 162.0 | 2708.1 | 59.8 | 615.0 | 12427.4 | 49.5 |
| Dyslipidaemia□ |  |  |  |  |  |  |  |  |  |  |  |  |
| Normal | 65.2 | 1985.5 | 32.8 | 213.2 | 5183.8 | 41.1 | 126.3 | 2503.5 | 50.5 | 404.7 | 9672.8 | 41.8 |
| Dyslipidaemic | 143.9 | 4282.8 | 33.6 | 323.8 | 5980.5 | 54.1 | 127.7 | 2298.4 | 55.6 | 595.3 | 12561.6 | 47.4 |
| Residential district |  |  |  |  |  |  |  |  |  |  |  |  |
| East | 45.8 | 1252.0 | 36.6 | 103.6 | 2285.8 | 45.3 | 62.5 | 1040.5 | 60.0 | 211.9 | 4578.3 | 46.3 |
| Tatsukawa | 27.5 | 1185.7 | 23.2 | 86.2 | 1778.0 | 48.5 | 37.3 | 783.9 | 47.6 | 151.0 | 3747.7 | 40.3 |
| South | 24.5 | 707.9 | 34.6 | 75.5 | 1434.9 | 52.6 | 26.3 | 556.9 | 47.1 | 126.2 | 2699.7 | 46.8 |
| Fudeoka | 28.6 | 795.0 | 36.0 | 56.3 | 1370.8 | 41.0 | 32.1 | 592.2 | 54.2 | 117.0 | 2758.0 | 42.4 |
| Central | 29.3 | 806.6 | 36.3 | 59.2 | 1359.0 | 43.5 | 34.2 | 523.2 | 65.3 | 122.6 | 2688.8 | 45.6 |
| Yoshiwara | 20.5 | 593.1 | 34.6 | 77.8 | 1247.8 | 62.3 | 22.2 | 433.0 | 51.3 | 120.5 | 2273.9 | 53.0 |
| West | 21.5 | 561.5 | 38.2 | 51.9 | 1078.0 | 48.1 | 22.4 | 493.5 | 45.3 | 95.7 | 2133.0 | 44.9 |
| Yogita | 11.3 | 366.4 | 30.9 | 26.7 | 610.0 | 43.8 | 17.1 | 378.7 | 45.2 | 55.2 | 1355.2 | 40.7 |
| HbA1c values |  |  |  |  |  |  |  |  |  |  |  |  |
| Mean (SD) | 5.51 | (0.43) |  | 5.46 | (0.41) |  | 5.41 | (0.41) |  | 5.46 | (0.42) |  |

Abbreviations: ALT, alanine transaminase; AST, aspartate aminotransferase; BMI, body mass index; HbA1c, haemoglobin A1C; IR, incidence rate†.
Results for multiple imputed variables (overweight or obesity‡, hypertension§, dyslipidaemia□, self-reported alcohol intake, self-reported smoking status and residential district) are averaged over 40 imputations.
†Incidence rate is reported per 1000 person-years.
‡Overweight or obesity is defined as a BMI ≥25 kg/m<sup>2</sup>.
§Hypertension is defined as systolic blood pressure ≥130 mmHg and/or diastolic blood pressure ≥80 mmHg.
□Dyslipidaemia is defined as serum low-density lipoprotein cholesterol ≥140 mg/dL, serum high-density lipoprotein cholesterol <40 mg/dL, and/or serum triglycerides ≥150 mg/dL.

**Table 2.** Descriptive statistics of all observations stratified by the AST/ALT ratio categories among 4395 non-diabetic Japanese female citizens of Zentsuji City (1998–2023)

|  | AST/ALT ratio classification |  |  |  |  |  |  |  |  | Total |  |  |
| --- | --- | --- | --- | --- | --- | --- | --- | --- | --- | --- | --- | --- |
|  | <1.0 |  |  | 1.0–<1.5 |  |  | ≥1.5 |  |  |  |  |  |
|  | Failure | PY | IR† | Failure | PY | IR† | Failure | PY | IR† | Failure | PY | IR† |
| Time at risk |  |  |  |  |  |  |  |  |  |  |  |  |
| Total |  | 5322.5 |  |  | 20052.3 |  |  | 11265.2 |  |  | 36639.9 |  |
| Minimum |  | 0.48 |  |  | 0.42 |  |  | 0.49 |  |  | 0.48 |  |
| Maximum |  | 21.3 |  |  | 24.5 |  |  | 24.6 |  |  | 24.7 |  |
| Mean |  | 4.09 |  |  | 6.07 |  |  | 4.99 |  |  | 8.34 |  |
| Median |  | 2.26 |  |  | 4.32 |  |  | 3.09 |  |  | 6.98 |  |
| Failure/IR† | 166 |  | 31.2 | 760 |  | 37.9 | 568 |  | 50.4 | 1494 |  | 40.8 |
| Variables |  |  |  |  |  |  |  |  |  |  |  |  |
| Age category in years |  |  |  |  |  |  |  |  |  |  |  |  |
| 34–59 | 67 | 2467.1 | 27.2 | 182 | 7566.3 | 24.1 | 97 | 3592.1 | 27.0 | 346 | 13625.5 | 25.4 |
| 60–69 | 60 | 2003.1 | 30.0 | 239 | 7015.2 | 34.1 | 126 | 3185.6 | 39.6 | 425 | 12203.9 | 34.8 |
| 70–100 | 39 | 852.3 | 45.8 | 339 | 5470.9 | 62.0 | 345 | 4487.4 | 76.9 | 723 | 10810.6 | 66.9 |
| BMI category‡ |  |  |  |  |  |  |  |  |  |  |  |  |
| Normal | 90 | 3113.2 | 28.9 | 527 | 15770.5 | 33.4 | 472 | 9848.6 | 47.9 | 1089 | 28732.3 | 37.9 |
| Overweight or obesity | 76 | 2209.3 | 34.4 | 233 | 4281.8 | 54.4 | 96 | 1416.6 | 67.8 | 405 | 7907.7 | 51.2 |
| Self-reported alcohol intake |  |  |  |  |  |  |  |  |  |  |  |  |
| Non- or seldom-drinker | 125.7 | 3977.9 | 31.6 | 609.5 | 14650.4 | 41.6 | 443.2 | 8586.9 | 51.6 | 1178.3 | 27215.2 | 43.3 |
| Drinker | 40.4 | 1344.6 | 30.0 | 150.6 | 5401.9 | 27.9 | 124.8 | 2678.2 | 46.6 | 315.7 | 9424.7 | 33.5 |
| Self-reported smoking status |  |  |  |  |  |  |  |  |  |  |  |  |
| Non- or ex-smoker | 161.2 | 4983.5 | 32.3 | 726.9 | 18986.2 | 38.3 | 553.5 | 10672.9 | 51.9 | 1441.5 | 34642.6 | 41.6 |
| Smoker | 4.8 | 339.0 | 14.2 | 33.2 | 1066.1 | 31.1 | 14.6 | 592.2 | 24.6 | 52.5 | 1997.4 | 26.3 |
| Hypertension§ |  |  |  |  |  |  |  |  |  |  |  |  |
| Normal | 62.0 | 2467.1 | 25.1 | 321.0 | 10513.5 | 30.5 | 225.0 | 5842.7 | 38.5 | 608.0 | 18823.4 | 32.3 |
| Hypertensive | 104.0 | 2855.4 | 36.4 | 439.0 | 9538.8 | 46.0 | 343.0 | 5422.4 | 63.3 | 886.0 | 17816.6 | 49.7 |
| Dyslipidaemia□ |  |  |  |  |  |  |  |  |  |  |  |  |
| Normal | 57.3 | 2009.0 | 28.5 | 315.5 | 9435.6 | 33.4 | 275.8 | 5806.2 | 47.5 | 648.6 | 17250.8 | 37.6 |
| Dyslipidaemic | 108.7 | 3313.5 | 32.8 | 444.5 | 10616.7 | 41.9 | 292.2 | 5458.9 | 53.5 | 845.4 | 19389.1 | 43.6 |
| Residential district |  |  |  |  |  |  |  |  |  |  |  |  |
| East | 39.7 | 1096.0 | 36.2 | 159.8 | 4041.9 | 39.5 | 109.0 | 2406.6 | 45.3 | 308.5 | 7544.5 | 40.9 |
| Tatsukawa | 27.5 | 821.1 | 33.5 | 111.2 | 3513.2 | 31.7 | 90.8 | 2052.7 | 44.2 | 229.5 | 6387.1 | 35.9 |
| South | 19.3 | 677.5 | 28.5 | 117.3 | 2789.8 | 42.0 | 94.7 | 1607.5 | 58.9 | 231.3 | 5074.9 | 45.6 |
| Fudeoka | 22.5 | 643.5 | 34.9 | 86.9 | 2542.8 | 34.2 | 65.7 | 1310.4 | 50.1 | 175.0 | 4496.7 | 38.9 |
| Central | 17.2 | 634.9 | 27.1 | 75.5 | 2008.7 | 37.6 | 57.5 | 1280.6 | 44.9 | 150.2 | 3924.3 | 38.3 |
| Yoshiwara | 16.3 | 590.6 | 27.6 | 88.5 | 2111.0 | 41.9 | 51.6 | 1032.6 | 49.9 | 156.4 | 3734.1 | 41.9 |
| West | 18.4 | 519.6 | 35.5 | 70.3 | 1999.9 | 35.2 | 61.6 | 939.1 | 65.6 | 150.3 | 3458.6 | 43.5 |
| Yogita | 5.1 | 339.2 | 15.1 | 50.7 | 1045.0 | 48.5 | 37.2 | 635.6 | 58.5 | 93.0 | 2019.8 | 46.0 |
| HbA1c values |  |  |  |  |  |  |  |  |  |  |  |  |
| Mean (SD) | 5.58 | (0.41) |  | 5.50 | (0.39) |  | 5.45 | (0.36) |  | 5.49 | (0.38) |  |

Abbreviations: ALT, alanine transaminase; AST, aspartate aminotransferase; BMI, body mass index; HbA1c, haemoglobin A1C; IR, incidence rate†.
Results for multiple imputed variables (overweight or obesity‡, hypertension§, dyslipidaemia□, self-reported alcohol intake, self-reported smoking status, and residential district) are averaged over 40 imputations.
†Incidence rate is reported per 1000 person-years.
‡Overweight or obesity is defined as a BMI ≥25 kg/m<sup>2</sup>.
§Hypertension is defined as systolic blood pressure ≥130 mmHg and/or diastolic blood pressure ≥80 mmHg.
□Dyslipidaemia is defined as serum low-density lipoprotein cholesterol ≥140 mg/dL, serum high-density lipoprotein cholesterol <40 mg/dL, and/or serum triglycerides ≥150 mg/dL.

Table 3 shows the estimation results for the relationship between AST/ALT and new-onset CKD in men and women. In the AST/ALT 1.0–<1.5 group, survival time to CKD onset was similar in both sexes and 4% shorter compared with the reference group, with all 95% CIs crossing 1 in all models. However, women with AST/ALT ≥1.5 showed a shorter survival time to CKD onset with a maximum of 9% (95% CI: 4%–14%), while men had a slightly shorter survival time to CKD onset by point estimates, but all 95% CIs crossed the null value.

**Table 3.** New onset of chronic kidney disease according to AST/ALT ratio categories among non-diabetic Japanese citizens of Zentsuji City, using the Weibull accelerated failure time model (1998–2023)

| AST/ALT ratio classification | PY | Failure | Crude | Model 1 | Model 2 | Model 3 | Model 4 |
| --- | --- | --- | --- | --- | --- | --- | --- |
|  |  |  | TR (95% CI) | aTR (95% CI) | aTR (95% CI) | aTR (95% CI) | aTR (95% CI) |
| **Men (n=2966)** |  |  |  |  |  |  |  |
| <1.0 (reference) | 6268.3 | 209 | 1.00 | 1.00 | 1.00 | 1.00 | 1.00 |
| 1.0–<1.5 | 11164.3 | 537 | 0.98 (0.95–1.02) | 0.98 (0.94–1.03) | 0.97 (0.93–1.01) | 0.96 (0.92–1.01) | 0.96 (0.92–1.01) |
| ≥1.5 | 4801.9 | 254 | 1.01 (0.96–1.05) | 1.00 (0.95–1.05) | 0.98 (0.93–1.03) | 0.98 (0.92–1.03) | 0.98 (0.92–1.03) |
| **Women (n=4395)** |  |  |  |  |  |  |  |
| <1.0 (reference) | 5322.5 | 166 | 1.00 | 1.00 | 1.00 | 1.00 | 1.00 |
| 1.0–<1.5 | 20052.3 | 760 | 0.98 (0.93–1.02) | 0.98 (0.94–1.02) | 0.96 (0.92–1.01) | 0.96 (0.91–1.00) | 0.96 (0.91–1.00) |
| ≥1.5 | 11265.2 | 568 | 0.93 (0.89–0.98) | 0.94 (0.90–0.99) | 0.92 (0.88–0.97) | 0.91 (0.86–0.96) | 0.91 (0.86–0.96) |

Abbreviations: ALT, alanine transaminase; AST, aspartate transferase; aTR, adjusted time ratio; BMI, body mass index; CI, confidence interval; HbA1c, haemoglobin A1C; PY, person-years; TR, time ratio.
†Overweight or obesity is defined as BMI ≥25 kg/m<sup>2</sup>.
‡Hypertension is defined as systolic blood pressure ≥130 mmHg and/or diastolic blood pressure ≥80 mmHg.
§Dyslipidaemia is defined as serum low-density lipoprotein cholesterol ≥140 mg/dL, serum high-density lipoprotein cholesterol <40 mg/dL and/or serum triglycerides ≥150 mg/dL.
Multiple imputed variables: overweight or obesity†, hypertension‡, dyslipidaemia§, self-reported alcohol intake, self-reported smoking status and residential district.
Model 1: Adjusted for age category (34–59 [reference]/60–69/70–100).
Model 2: Adjusted for the variable of Model 1, BMI category† (normal [reference]/overweight or obesity), self-reported alcohol intake (non- or seldom-drinker [reference]/yes), and self-reported smoking status (non- or ex-smoker [reference]/smoker).
Model 3: Adjusted for all variables of Model 2, hypertension‡ (normal [reference]/hypertensive), dyslipidaemia§ (normal [reference]/dyslipidaemic), and HbA1c values.
Model 4: Adjusted for all variables of Model 3 and residential district (East [reference]/Tatsukawa/South/Fudeoka/Central/Yoshikawa/West/Yogita).

### Sensitivity analyses

The first sensitivity analysis used ALT, AST, and GGT as the exposure variables. Table S1 shows the results using ALT as the exposure variable. Women had higher ALT than men, with ALT ≥40 U/L accounting for 30.7% of the total follow-up time (8.90% in men). Both sexes had longer survival times with higher ALT, with all 95% CIs crossing 1 in all models. Men had slightly higher point estimates of survival time than women in the elevated ALT categories.

Table S2 presents the results with AST as the exposure variable. Men had slightly higher AST than women, with AST ≥30 U/L accounting for 19.2% of the total follow-up time (9.62% in women). The trends in the estimation results differed between men and women. In men, higher AST was associated with longer survival time to CKD onset by point estimates, while in women, higher AST was associated with slightly shorter survival time to CKD onset by point estimates. Regardless of sex, all 95% CIs crossed 1 in all models.

Table S3 shows the findings when GGT was the exposure variable. Men had higher GGT than women, with GGT ≥30 U/L accounting for 49.7% of the total follow-up time (19.2% in women). After adjusting for all covariates including alcohol intake, all point estimates of survival were around the null value for both sexes, with all 95% CIs crossing 1. There was no clear evidence for a dose-response relationship in either sex.

Table S4 shows the results of the second sensitivity analysis using the stringent definition of CKD, in which two consecutive observations of eGFR <60 mL/min/1.73 m^2^ were regarded as CKD. The men and women excluded for prevalent CKD at study entry comprised 3.56% and 8.88% in the 45–59 age group, 15.9% and 21.2% in the 60–69 age group, and 27.7% and 33.5% in the 70–100 age group, respectively. In total, fewer participants developed CKD (19.3% of men and 19.8% of women) compared with the main analysis (Tables 1 and 2). The mean ages of the participants at CKD onset were 70.6 years for men and 69.3 years for women, being slightly older than the main analysis. The characteristics of both sexes were similar to the main analysis. Both sexes showed attenuated results with longer survival times to CKD onset and 95% CIs crossing 1, similar to the full model (Table 3).

Table S5 presents the results of the third sensitivity analysis using dipstick proteinuria ≥1+ as the definition of CKD. All AST/ALT categories had a longer follow-up period regardless of sex compared with the main analysis (Tables 1 and 2). Meanwhile, the proportions of incident CKD were lower than those in the main analysis for both sexes. Survival to incident CKD was slightly longer compared with the main analysis for both men and women, with all 95% CIs crossing the null value.

Finally, Table S6 shows the results of the Cox PH model. In both sexes, the hazard ratio estimates tended to be >1, although most of them were not statistically significant. These trends were similar to those in the main analysis using the Weibull AFT model (Table 3).

## Discussion

### Main analysis

After approximately 25 years of follow-up, a dose-response relationship between AST/ALT categories and subsequent CKD development was only found in women, with a 9% shorter survival time to CKD onset for AST/ALT ≥1.5 compared with AST/ALT <1.0. A previous cross-sectional study involving 29133 middle-aged Japanese women with and without diabetes found that AST/ALT <1 had a 1.43 times stronger association (95% CI: 1.30–1.58) with prevalent CKD than AST/ALT ≥1, after adjustment for all covariates including diabetes[9]. The trend in the present study contradicted the results of the previous study.

The discrepancy may mainly arise through differences in the study designs and participant CKD severities. The present study was longitudinal, whereas the previous study was cross-sectional—a design often used to infer causality before follow-up studies[9,28]. We followed participants without CKD at study entry to see whether they developed CKD, and >95% of the participants who did were in stage G3a, the mildest stage. In contrast, the previous cross-sectional study included participants with and without CKD at any stage, from G3a to G5[9]. Other cross-sectional studies showed that liver enzyme levels decrease with advancing CKD stages, even in haemodialysis patients with end-stage renal disease, regardless of viral hepatitis[29–31]. CKD progression damages hepatocytes via uremic toxins and chronic oxidative stress, potentially lowering AST/ALT, and CKD may involve an initial increase in AST/ALT[32]. Therefore, the pathway between AST/ALT and CKD may involve an initial increase in AST/ALT affecting the onset of CKD, as observed in the present study, followed by an AST/ALT decrease as CKD worsens.

Another reason for the discrepancy may be that the present study only included non-diabetic participants, while the previous Japanese cross-sectional study included both diabetic and non-diabetic participants[9], which may have influenced the results. In an Asian cohort study involving 87883 Chinese adults, a one-unit increase in AST/ALT was associated with a 0.56-fold risk of developing diabetes (95% CI: 0.37–0.85) after adjusting for all covariates including fasting plasma glucose and family history of diabetes[33]. In the previous Japanese cross-sectional study, participants with CKD tended to have diabetes of any severity (7.2% of CKD participants) compared with those without CKD (3.4% of non-CKD participants)[9]. This suggests that even after controlling diabetes, there may have been residual confounding between AST/ALT and new-onset CKD, which may have influenced the reversed trend observed in the previous studies[9,11,33].

In a study on people in the Japanese general population who underwent health check-ups in 11 prefectures, the prevalences of CKD (eGFR <60 mL/min/1.73 m^2^) in men and women by age groups were 3.97% and 2.57% in their 40s, 7.48% and 6.82% in their 50s, 15.8% and 14.8% in their 60s, 27.7% and 31.8% in their 70s, and 44.6% and 46.1% in their 80s and above, respectively[34]. The fact that the maximum age of the participants in the present study was very old (100 years) may partly explain the differences between the populations.

### Sensitivity analyses

In the first sensitivity analysis, AST, ALT, and GGT were used as the exposure variables (Tables S1–S3). In the present study, elevated ALT and GGT showed a slightly longer survival time to CKD onset in both sexes by point estimates, but all 95% CIs crossed 1 in all models. In the previous cross-sectional study from Japan, elevated ALT >40 U/L and GGT >50 U/L were 1.53 (95% CI: 1.31–1.79) and 1.60 (95% CI: 1.42–1.80) times more strongly associated with CKD prevalence than ALT ≤40 U/L and GGT ≤50 U/L, respectively, after full adjustment for confounders including diabetes (no data were provided for AST)[9]. To our knowledge, no follow-up studies involving non-diabetic Asian participants have examined ALT or AST and subsequent development of CKD. However, one Asian cohort study did investigate the relationship between GGT and CKD onset among 9341 non-diabetic Japanese men in the Kansai Healthcare Study[23]. In that cohort study, higher GGT showed no clear association with CKD onset in the non-diabetic Japanese participants, consistent with the trend in the present study. In summary, ALT, AST, and GGT are not risk factors for CKD development in Japanese adults without diabetes. These findings imply that AST/ALT may reflect liver function more sensitively than the individual levels of AST and ALT. If liver diseases are considered to influence incident CKD, our findings suggest the clinical relevance of using AST/ALT for assessment. Further research is warranted to explore this hypothesis.

The results of the second sensitivity analysis using a stringent definition of CKD showed that fewer participants were excluded due to CKD prevalence at study entry, and fewer participants developed CKD after follow-up at a slightly older age and with higher point estimates than the main analysis (Table S4). The present study used the results of annual health check-ups. Therefore, the definition in the second sensitivity analysis was more stringent than the KDIGO definition of CKD, which is renal dysfunction for >3 months[7]. The stringent definition minimized the visit-to-visit variability of eGFR to define CKD. However, all participants who developed CKD at the last observation were classified as non-CKD, possibly underestimating the results based on the overly stringent definition of CKD.

In the third sensitivity analysis using dipstick proteinuria ≥1+ as the CKD definition, the proportion of CKD cases decreased, suggesting that use of dipstick proteinuria alone to define CKD may miss some CKD cases and lead to an underestimation of the results. These findings are consistent with a cross-sectional study on the general Japanese population (n=538846), which found that 14.4% had eGFR <60 mL/min/1.73 m^2^, 5.2% had dipstick proteinuria ≥1+, and 18.1% had either or both[35].

Finally, in the fourth sensitivity analysis, most hazard ratio estimates were >1. These findings are consistent with the results of the Weibull AFT model, which indicated a shorter survival time to development of CKD.

### Strengths and limitations

The strengths of the study are the single ethnicity of the participants, the sufficiently long follow-up period for detection of CKD development, and the sex-stratified analysis, given that the distribution of liver enzymes varies by sex[1]. To the best of our knowledge, this is the first sex-stratified longitudinal study evaluating AST/ALT and new-onset CKD in a non-diabetic Asian population.

Several limitations of the study should also be noted. First, we used health check-up data in a single city, with the participants themselves deciding whether or not to undergo the health check-ups. Thus, the participants in the study were not randomly sampled from the entire Japanese population and were not representative of Japan as a whole. Thus, the present results may not be generalizable to other populations.

Second, the study participants were healthier than the general population because they continued to undergo health check-ups over a long period. Unhealthy participants may have tended to be right-censored during the follow-up period because of competing risks such as serious illnesses requiring regular medical visits, hospitalization, or death (data unavailable). Therefore, there is a possibility that unhealthy participants were more likely to be dropped from the analysis during the follow-up (e.g., death would have prevented them from receiving further checkups, leading to their exclusion from the cohort). This could result in an underestimation of the study results. Furthermore, because of the long follow-up period in the study, a built-in selection bias may have existed, leading to further underestimation of the results[36].

Third, there is a possibility of misclassification of renal outcomes. In this study, only SCr and dipstick proteinuria were measured, and thus the albumin-to-creatinine ratio (ACR) and protein-to-creatinine ratio (PCR) were unavailable, although they are recommended for definition and staging of CKD[7,37]. A previous cross-sectional study on Japanese workers (n=5383) reported that the dipstick urine test had high specificity but low sensitivity for detecting ACR albuminuria and PCR proteinuria[38], and therefore the results of the present study may be underestimated. To define CKD, we used SCr, which is affected by higher muscle mass and dehydration[39,40], for calculation of eGFR. Previous studies reported that men have higher skeletal mass than women, that skeletal mass decreases with age, and that dehydration is more common in older people[41,42]. In the present study, the results were described separately for men and women, with 72.2% of the men in the entire follow-up period being ≥60 years of age (62.8% in women). Thus, due to the small number of young participants, the effect of high muscle mass should be negligible. Nevertheless, there may be residual bias derived from older participants even after controlling for age, and the findings may have been overestimated. However, in the present analysis, we used the three-variable Japanese equation, which performs better than conventional equations for calculation of eGFR, and thus the effect of age may have been smaller[16].

Fourth, there are several unmeasured confounding factors. For example, the prevalence and treatment of liver diseases are unknown. Since liver enzymes may increase or decrease depending on the types of liver diseases, our findings could be overestimated or underestimated. Furthermore, our results may be overestimated due to confounding bias from medication use[11,43–45]. Moreover, information on hepatitis B and C virus infection was unavailable, and patients with hepatitis B or C tend to have AST/ALT <1 and be more likely to develop CKD[1,46,47]. However, the prevalences of hepatitis B and C in the Japanese population are quite low (3.0% and 0.13%, respectively)[48,49], suggesting that there may have been little impact on the study results.

Finally, the study used interval data, for which the observation intervals depended on the participants, and thus the actual date of CKD onset was unknown.

## Conclusions

In the present study using administrative health check-up data for healthy middle-aged and older Japanese without diabetes, there was a dose-response relationship between AST/ALT and subsequent development of CKD in women (higher AST/ALT was associated with shorter survival time to CKD onset), but no clear trend in men. The present findings suggest that AST/ALT values commonly obtained in daily practice can be used as an indicator for early detection of CKD in non-diabetic Japanese women.

## Data Availability

All data relevant to this study are included in the article and supplemental files.

## Ethics

We used data anonymized before receipt. The Ethics Committee of Okayama University Graduate School of Medicine, Dentistry and Pharmaceutical Sciences and Okayama University Hospital approved this study (No. K1708-040). The Ethics committee waived the need for informed consent. This study was conducted in accordance with the Declaration of Helsinki and Japanese Ethical Guidelines for Medical and Biological Research Involving Human Subjects.

## Disclosure of potential conflicts of interest

Employment: Yukari Okawa (Zentsuji City). Toshiharu Mitsuhashi and Etsuji Suzuki: no competing interests to declare. This research is self-funded and received no specific grant from funding agencies in the public, commercial, or not-for-profit sectors.

## Acknowledgements

We appreciate all participants of this study, Ayaka Nakatsu, Masako Matsumoto, Mayumi Kitadani and all local government officers of Zentsuji City for their support and contribution. The authors thank Alison Sherwin, PhD, from Edanz (https://jp.edanz.com/ac) for editing a draft of this manuscript.

## Data accessibility statement

All data relevant to this study are included in the article and supplemental files.

## Author contributions

**Etsuji Suzuki:** Conceptualization, Methodology, Writing - review & editing, Supervision. **Toshiharu Mitsuhashi:** Conceptualization, Methodology, Writing - review & editing, Supervision. **Yukari Okawa:** Conceptualization, Methodology, Formal analysis, Investigation, Data curation, Writing - original draft, Writing - review & editing, Visualization, Project administration.

## Notes

### Summary of Updates

We submitted the wrong file (manuscript) in the previous version.

