## Supplementary Tables for "Aspartate/alanine aminotransferase ratio and development of chronic kidney disease in non-diabetic men and women: a population-based longitudinal study in Kagawa, Japan"

**Table S1.** New onset of chronic kidney disease according to ALT categories among non-diabetic Japanese citizens of Zentsuji City, using the Weibull accelerated failure time model (1998–2023)

| | | | Crude | Model 1 | Model 2 | Model 3 | Model 4 |
|---|---|---|---|---|---|---|---|
| ALT classification | PY | Failure | TR (95% CI) | aTR (95% CI) | aTR (95% CI) | aTR (95% CI) | aTR (95% CI) |
| **Men (n=2966)** | | | | | | | |
| <30 U/L (reference) | 18065.1 | 861 | 1.00 | 1.00 | 1.00 | 1.00 | 1.00 |
| 30–<40 U/L | 2191.4 | 79 | 1.02 (0.97–1.08) | 1.03 (0.97–1.09) | 1.04 (0.98–1.11) | 1.05 (0.99–1.12) | 1.05 (0.98–1.11) |
| ≥40 U/L | 1977.9 | 60 | 1.03 (0.97–1.10) | 1.03 (0.96–1.11) | 1.05 (0.98–1.13) | 1.06 (0.99–1.14) | 1.06 (0.99–1.14) |
| **Women (n=4395)** | | | | | | | |
| <30 U/L (reference) | 5322.5 | 166 | 1.00 | 1.00 | 1.00 | 1.00 | 1.00 |
| 30–<40 U/L | 20052.3 | 760 | 1.01 (0.95–1.08) | 1.01 (0.95–1.07) | 1.02 (0.96–1.08) | 1.03 (0.96–1.09) | 1.03 (0.96–1.09) |
| ≥40 U/L | 11265.2 | 568 | 1.00 (0.93–1.08) | 1.00 (0.93–1.07) | 1.02 (0.95–1.10) | 1.03 (0.95–1.11) | 1.03 (0.95–1.11) |

Abbreviations: ALT, alanine transaminase; aTR, adjusted time ratio; BMI, body mass index; CI, confidence interval; HbA1c, haemoglobin A1C; PY, person-years; TR, time ratio.

†Overweight or obesity is defined as BMI ≥25 kg/m$^2$.

‡Hypertension is defined as systolic blood pressure ≥130 mmHg and/or diastolic blood pressure ≥80 mmHg.

§Dyslipidaemia is defined as serum low-density lipoprotein cholesterol ≥140 mg/dL, serum high-density lipoprotein cholesterol <40 mg/dL, and/or serum triglycerides ≥150 mg/dL.

Multiple imputed variables: overweight or obesity†, hypertension‡, dyslipidaemia§, self-reported alcohol intake, self-reported smoking status, and residential district.

Model 1: Adjusted for age category (34–59 [reference]/60–69/70–100).

Model 2: Adjusted for the variable of Model 1, BMI category† (normal [reference]/overweight or obesity), self-reported alcohol intake (non- or seldom-drinker [reference]/drinker), and self-reported smoking status (non- or ex-smoker [reference]/smoker).

Model 3: Adjusted for all variables of Model 2, hypertension‡ (normal [reference]/hypertensive), dyslipidaemia§ (normal [reference]/dyslipidaemic), and HbA1c values.

Model 4: Adjusted for all variables of Model 3 and residential district (East [reference]/Tatsukawa/South/Fudeoka/Central/Yoshikawa/West/Yogita).

**Table S2**. New onset of chronic kidney disease according to the AST categories among non-diabetic Japanese citizens of Zentsuji City, using the Weibull accelerated failure time model (1998–2023)

| | | | Crude | Model 1 | Model 2 | Model 3 | Model 4 |
|---|---|---|---|---|---|---|---|
| AST classification | PY | Failure | TR (95% CI) | aTR (95% CI) | aTR (95% CI) | aTR (95% CI) | aTR (95% CI) |
| **Men (n=2966)** | | | | | | | |
| <30 U/L (reference) | 17965.7 | 812 | 1.00 | 1.00 | 1.00 | 1.00 | 1.00 |
| 30–<40 U/L | 2878.4 | 138 | 0.99 (0.95–1.04) | 0.99 (0.95–1.04) | 1.00 (0.95–1.04) | 1.00 (0.96–1.05) | 1.00 (0.96–1.05) |
| ≥40 U/L | 1390.4 | 50 | 1.04 (0.97–1.11) | 1.04 (0.97–1.12) | 1.05 (0.97–1.13) | 1.06 (0.98–1.14) | 1.06 (0.98–1.14) |
| **Women (n=4395)** | | | | | | | |
| <30 U/L (reference) | 33113.8 | 1318 | 1.00 | 1.00 | 1.00 | 1.00 | 1.00 |
| 30–<40 U/L | 2500.5 | 125 | 0.98 (0.93–1.03) | 0.98 (0.94–1.03) | 0.98 (0.94–1.03) | 0.98 (0.94–1.03) | 0.98 (0.94–1.03) |
| ≥40 U/L | 1025.6 | 51 | 0.96 (0.90–1.04) | 0.97 (0.90–1.03) | 0.98 (0.91–1.05) | 0.98 (0.91–1.05) | 0.98 (0.91–1.05) |

Abbreviations: AST, aspartate transferase; aTR, adjusted time ratio; BMI, body mass index; CI, confidence interval; HbA1c, haemoglobin A1C; PY, person-years; TR, time ratio.

†Overweight or obesity is defined as BMI ≥25 kg/$m^2$.

‡Hypertension is defined as systolic blood pressure ≥130 mmHg and/or diastolic blood pressure ≥80 mmHg.

§Dyslipidaemia is defined as serum low-density lipoprotein cholesterol ≥140 mg/dL, serum high-density lipoprotein cholesterol <40 mg/dL, and/or serum triglycerides ≥150 mg/dL.

Multiple imputed variables: overweight or obesity†, hypertension‡, dyslipidaemia§, self-reported alcohol intake, self-reported smoking status, and residential district.

Model 1: Adjusted for age category (34–59 [reference]/60–69/70–100).

Model 2: Adjusted for the variable of Model 1, BMI category† (normal [reference]/overweight or obesity), self-reported alcohol intake (non- or seldom-drinker [reference]/drinker), and self-reported smoking status (non- or ex-smoker [reference]/smoker).

Model 3: Adjusted for all variables of Model 2, hypertension‡ (normal [reference]/hypertensive), dyslipidaemia§ (normal [reference]/dyslipidaemic), and HbA1c values.

Model 4: Adjusted for all variables of Model 3 and residential district (East [reference]/Tatsukawa/South/Fudeoka/Central/Yoshikawa/West/Yogita).

**Table S3**. New onset of chronic kidney disease according to the GGT categories among non-diabetic Japanese citizens of Zentsuji City, using the Weibull accelerated failure time model (1998–2023)

| | | | Crude | Model 1 | Model 2 | Model 3 | Model 4 |
|---|---|---|---|---|---|---|---|
| GGT classification | PY | Failure | TR (95% CI) | aTR (95% CI) | aTR (95% CI) | aTR (95% CI) | aTR (95% CI) |
| **Men (n=2966)** | | | | | | | |
| <30 U/L (reference) | 11175.4 | 539 | 1.00 | 1.00 | 1.00 | 1.00 | 1.00 |
| 30–<50 U/L | 5720.4 | 250 | 1.00 (0.96–1.03) | 1.00 (0.96–1.04) | 1.01 (0.97–1.05) | 1.01 (0.97–1.05) | 1.01 (0.97–1.05) |
| ≥50 U/L | 5338.7 | 211 | 0.99 (0.95–1.03) | 0.99 (0.95–1.03) | 1.00 (0.96–1.04) | 1.01 (0.96–1.05) | 1.01 (0.96–1.05) |
| **Women (n=4395)** | | | | | | | |
| <30 U/L (reference) | 29615.7 | 1226 | 1.00 | 1.00 | 1.00 | 1.00 | 1.00 |
| 30–<50 U/L | 4521.5 | 172 | 1.03 (0.99–1.07) | 1.02 (0.98–1.06) | 1.03 (0.99–1.07) | 1.03 (0.99–1.08) | 1.03 (0.99–1.08) |
| ≥50 U/L | 2502.8 | 96 | 1.01 (0.95–1.06) | 1.00 (0.95–1.06) | 1.01 (0.96–1.06) | 1.02 (0.96–1.07) | 1.01 (0.96–1.07) |

Abbreviations: aTR, adjusted time ratio; BMI, body mass index; CI, confidence interval; GGT, gamma-glutamyltransferase; HbA1c, haemoglobin A1C; PY, person-years; TR, time ratio.

†Overweight or obesity is defined as BMI ≥25 kg/m$^2$.

‡Hypertension is defined as systolic blood pressure ≥130 mmHg and/or diastolic blood pressure ≥80 mmHg.

§Dyslipidaemia is defined as serum low-density lipoprotein cholesterol ≥140 mg/dL, serum high-density lipoprotein cholesterol <40 mg/dL and/or serum triglycerides ≥150 mg/dL.

Multiple imputed variables: overweight or obesity†, hypertension‡, dyslipidaemia§, self-reported alcohol intake, self-reported smoking status, and residential district.

Model 1: Adjusted for age category (34–59 [reference]/60–69/70–100).

Model 2: Adjusted for the variable of Model 1, BMI category† (normal [reference]/overweight or obesity), self-reported alcohol intake (non- or seldom-drinker [reference]/drinker), and self-reported smoking status (non- or ex-smoker [reference]/smoker).

Model 3: Adjusted for all variables of Model 2, hypertension‡ (normal [reference]/hypertensive), dyslipidaemia§ (normal [reference]/dyslipidaemic), and HbA1c values.

Model 4: Adjusted for all variables of Model 3 and residential district (East [reference]/Tatsukawa/South/Fudeoka/Central/Yoshikawa/West/Yogita).

**Table S4.** New onset of chronic kidney disease according to AST/ALT ratio categories among non-diabetic Japanese citizens of Zentsuji City (1998–2023) using the Weibull accelerated failure time model with the stringent definition of CKD, in which two consecutive observations of eGFR <60 mL/min/1.73 $m^2$ were considered CKD (1998–2023)

| | | | Crude | Model 1 | Model 2 | Model 3 | Model 4 |
|---|---|---|---|---|---|---|---|
| AST/ALT ratio classification | PY | Failure | TR (95% CI) | aTR (95% CI) | aTR (95% CI) | aTR (95% CI) | aTR (95% CI) |
| **Men (n=3270)** | | | | | | | |
| <1.0 (reference) | 7228.1 | 125 | 1.00 | 1.00 | 1.00 | 1.00 | 1.00 |
| 1.0–<1.5 | 13654.0 | 350 | 0.99 (0.95–1.03) | 0.99 (0.95–1.03) | 0.98 (0.94–1.02) | 0.98 (0.94–1.02) | 0.98 (0.94–1.02) |
| ≥1.5 | 6192.9 | 156 | 1.04 (0.99–1.09) | 1.03 (0.98–1.08) | 1.01 (0.97–1.06) | 1.01 (0.96–1.05) | 1.01 (0.96–1.05) |
| **Women (n=5013)** | | | | | | | |
| <1.0 (reference) | 6572.0 | 91 | 1.00 | 1.00 | 1.00 | 1.00 | 1.00 |
| 1.0–<1.5 | 25547.3 | 500 | 0.95 (0.90–1.00) | 0.96 (0.91–1.01) | 1.00 (0.93–1.06) | 0.99 (0.93–1.06) | 0.99 (0.92–1.06) |
| ≥1.5 | 15494.4 | 400 | 0.93 (0.88–0.98) | 0.94 (0.88–0.99) | 0.96 (0.90–1.02) | 0.94 (0.88–1.01) | 0.94 (0.88–1.01) |

Abbreviations: ALT, alanine transaminase; AST, aspartate transferase; aTR, adjusted time ratio; BMI, body mass index; CI, confidence interval; HbA1c, haemoglobin A1C; PY, person-years; TR, time ratio.

†Overweight or obesity is defined as BMI ≥25 kg/$m^2$.

‡Hypertension is defined as systolic blood pressure ≥130 mmHg and/or diastolic blood pressure ≥80 mmHg.

§Dyslipidaemia is defined as serum low-density lipoprotein cholesterol ≥140 mg/dL, serum high-density lipoprotein cholesterol <40 mg/dL, and/or serum triglycerides ≥150 mg/dL.

Multiple imputed variables: overweight or obesity†, hypertension‡, dyslipidaemia§, self-reported alcohol intake, self-reported smoking status, and residential district.

Model 1: Adjusted for age category (34–59 [reference]/60–69/70–100).

Model 2: Adjusted for the variable of Model 1, BMI category† (normal [reference]/overweight or obesity), self-reported alcohol intake (non- or seldom-drinker [reference]/drinker), and self-reported smoking status (non- or ex-smoker [reference]/smoker). A multiplicative term (AST/ALT ratio classification × overweight or obesity†) was added for men.

Model 3: Adjusted for all variables of Model 2, hypertension‡ (normal [reference]/hypertensive), dyslipidaemia§ (normal [reference]/dyslipidaemic), and HbA1c values. A multiplicative term (AST/ALT ratio classification × overweight or obesity†) was added for men.

Model 4: Adjusted for all variables of Model 3 and residential district (East [reference]/Tatsukawa/South/Fudeoka/Central/Yoshikawa/West/Yogita). A multiplicative term (AST/ALT ratio classification × overweight or obesity†) was added for men.

**Table S5**. New onset of chronic kidney disease according to the AST/ALT ratio classification among non-diabetic Japanese citizens of Zentsuji City, with proteinuria ≥1+ was considered CKD, using the Weibull accelerated failure time model (1998–2023)

| | | | Crude | Model 1 | Model 2 | Model 3 | Model 4 |
|---|---|---|---|---|---|---|---|
| AST/ALT ratio classification | PY | Failure | TR (95% CI) | aTR (95% CI) | aTR (95% CI) | aTR (95% CI) | aTR (95% CI) |
| **Men (n=3957)** | | | | | | | |
| <1.0 (reference) | 8441.5 | 161 | 1.00 | 1.00 | 1.00 | 1.00 | 1.00 |
| 1.0–<1.5 | 17108.6 | 354 | 1.04 (0.99–1.10) | 1.03 (0.99–1.07) | 1.02 (0.98–1.06) | 1.02 (0.97–1.06) | 1.02 (0.98–1.06) |
| ≥1.5 | 8321.8 | 217 | 1.02 (0.95–1.08) | 1.02 (0.97–1.06) | 1.00 (0.96–1.06) | 1.00 (0.95–1.05) | 1.00 (0.95–1.05) |
| **Women (n=6458)** | | | | | | | |
| <1.0 (reference) | 7997.3 | 119 | 1.00 | 1.00 | 1.00 | 1.00 | 1.00 |
| 1.0–<1.5 | 33578.2 | 456 | 1.06 (1.01–1.11) | 1.06 (1.01–1.11) | 1.04 (0.99–1.09) | 1.04 (0.99–1.09) | 1.04 (0.99–1.09) |
| ≥1.5 | 23766.2 | 478 | 1.02 (0.97–1.07) | 1.02 (0.98–1.07) | 1.00 (0.95–1.05) | 1.00 (0.95–1.05) | 1.00 (0.95–1.05) |

Abbreviations: ALT, alanine transaminase; AST, aspartate transferase; aTR, adjusted time ratio; BMI, body mass index; CI, confidence interval; HbA1c, hemoglobin A1C; PY, person-years; TR, time ratio.

†Overweight or obesity is defined as BMI ≥25 kg/m$^2$.

‡Hypertension is defined as systolic blood pressure ≥130 mmHg and/or diastolic blood pressure ≥80 mmHg.

§Dyslipidemia is defined as serum low-density lipoprotein cholesterol ≥140 mg/dL, serum high-density lipoprotein cholesterol <40 mg/dL, and/or serum triglycerides ≥150 mg/dL.

Multiple imputed variables: overweight or obesity†, hypertension‡, dyslipidemia§, self-reported drinking status, self-reported smoking status, and residential district.

Model 1: Adjusted for age category (34–59 [reference]/60–69/≥70).

Model 2: Adjusted for the variable of Model 1, BMI category† (normal weight [reference]/overweight or obesity), self-reported drinking status (nondrinker [reference]/drinker), and self-reported smoking status (nonsmoker [reference]/smoker).

Model 3: Adjusted for all variables of Model 2, hypertension‡ (no [reference]/yes), dyslipidemia§ (no [reference]/yes), and HbA1c values.

Model 4: Adjusted for all variables of Model 3 and residential district (East [reference]/Tatsukawa/South/Fudeoka/Central/Yoshikawa/West/Yogita).

**Table S6**. New onset of chronic kidney disease according to the AST/ALT ratio classification among non-diabetic Japanese citizens of Zentsuji City, using the Cox proportional hazards model (1998–2023)

| | | | Crude | Model 1 | Model 2 | Model 3 | Model 4 |
|---|---|---|---|---|---|---|---|
| AST/ALT ratio classification | PY | Failure | HR (95% CI) | aHR (95% CI) | aHR (95% CI) | aHR (95% CI) | aHR (95% CI) |
| **Men (n=2966)** | | | | | | | |
| <1.0 (reference) | 6268.3 | 209 | 1.00 | 1.00 | 1.00 | 1.00 | 1.00 |
| 1.0–<1.5 | 11164.3 | 537 | 1.08 (0.92–1.28) | 1.09 (0.92–1.29) | 1.15 (0.97–1.36) | 1.18 (0.99–1.40) | 1.17 (0.99–1.39) |
| ≥1.5 | 4801.9 | 254 | 0.99 (0.81–1.20) | 0.99 (0.82–1.21) | 1.09 (0.89–1.33) | 1.11 (0.91–1.37) | 1.11 (0.91–1.37) |
| **Women (n=4395)** | | | | | | | |
| <1.0 (reference) | 5322.5 | 166 | 1.00 | 1.00 | 1.00 | 1.00 | 1.00 |
| 1.0–<1.5 | 20052.3 | 760 | 1.05 (0.89–1.25) | 1.06 (0.89–1.25) | 1.12 (0.94–1.34) | 1.16 (0.97–1.39) | 1.16 (0.97–1.39) |
| ≥1.5 | 11265.2 | 568 | 1.18 (0.99–1.42) | 1.19 (0.99–1.43) | 1.30 (1.08–1.58) | 1.36 (1.13–1.65) | 1.37 (1.13–1.66) |

Abbreviations: ALT, alanine transaminase; AST, aspartate transferase; aHR, adjusted hazard ratio; aTR, adjusted time ratio; BMI, body mass index; CI, confidence interval; HbA1c, hemoglobin A1C; HR, hazard ratio; PY, person-years; TR, time ratio.

†Overweight or obesity is defined as BMI ≥25 kg/m$^2$.

‡Hypertension is defined as systolic blood pressure ≥130 mmHg and/or diastolic blood pressure ≥80 mmHg.

§Dyslipidemia is defined as serum low-density lipoprotein cholesterol ≥140 mg/dL, serum high-density lipoprotein cholesterol <40 mg/dL, and/or serum triglycerides ≥150 mg/dL.

Multiple imputed variables: overweight or obesity†, hypertension‡, dyslipidemia§, self-reported drinking status, self-reported smoking status, and residential district.

Model 1: Adjusted for age category (34–59 [reference]/60–69/≥70).

Model 2: Adjusted for the variable of Model 1, BMI category† (normal weight [reference]/overweight or obesity), self-reported drinking status (nondrinker [reference]/drinker), and self-reported smoking status (nonsmoker [reference]/smoker).

Model 3: Adjusted for all variables of Model 2, hypertension‡ (no [reference]/yes), dyslipidemia§ (no [reference]/yes), and HbA1c values.

Model 4: Adjusted for all variables of Model 3 and residential district (East [reference]/Tatsukawa/South/Fudeoka/Central/Yoshikawa/West/Yogita).
